# Multi-region sampling of the human small intestine using an ingestible device

**DOI:** 10.64898/2026.06.09.26353912

**Authors:** Beverly Fu, Leila Bronnert DeSchepper, Jiawei Sun, Saria A. McKeithen-Mead, Bennett Kapili, Pablo Ochoa-Andersen, Sean Paul Spencer, Touran Fardeen, Maya Ricardo, Vanessa El Kamari, Sidhartha Sinha, David A. Relman, Jessica A. Grembi, Dari Shalon, Sylvie Estrela, Kerwyn Casey Huang

## Abstract

The human small intestine (SI) plays a central role in nutrient processing, host–microbe interactions, and immune regulation, yet remains poorly characterized due to the lack of minimally disruptive sampling methods. Here, we present a protocol for deploying, recovering, and analyzing samples collected using an ingestible device that enables multi-region, lumen-targeted SI sampling during normal digestion. The device incorporates a ∼30-cm collapsible tube wound into pH- or time-responsive layers that sequentially unfurl *in situ*, typically capturing three spatially ordered samples with high yield and reliable retrieval. This protocol outlines study design, participant handling, device recovery, contamination control, and standardized workflows for analyses, including cell quantification, culturomics, sequencing, and metabolomics. We further describe benchmarking approaches for evaluating spatial resolution and strategies for assay prioritization when sample volume is limiting. By reducing participant burden and facilitating integration with stool, saliva, and clinical metadata, this approach enables longitudinal and large-cohort studies linking SI microbial ecology and host physiology to human health.

## Introduction

The small intestine (SI) is central to human physiology serving as the primary site of nutrient digestion and absorption, immune training, and metabolic regulation^1^. Disruption of the SI environment has been implicated in a broad spectrum of diseases, including malnutrition^2^, inflammatory bowel disease (IBD)^3^, and metabolic disorders^3^. Despite its biological importance, our understanding of how SI-resident microbial communities and their metabolites influence host physiology and shape human health remains limited.

Existing approaches for interrogating the SI have substantial scientific and practical constraints. Stool sampling, although widely used, does not reflect small-intestinal processes^4^. Endoscopy and biopsy-based procedures provide direct tissue access^5^, including mucosa-associated communities, but require fasting and procedural sedation or anesthesia, potentially altering the luminal environment at the time of sampling. Naso-or oro-intestinal aspiration catheter-based methods permit luminal collection, but are invasive, low-throughput, and susceptible to contamination^6^. Continuous sampling in individuals with SI stomas offers direct luminal access, although altered anatomy and physiology may influence microbiome composition and local immune states^7^.

Large-scale organ donor studies have revealed the cellular and transcriptional complexity of the SI microbiome^8,9^; however, these investigations are inherently cross-sectional and often represent perturbed states due to medical intervention. Collectively, these limitations have constrained efforts to characterize spatial gradients, dynamic responses, and longitudinal variation within the human SI under normal digestive conditions, motivating the development of minimally disruptive technologies capable of multi-region, longitudinal sampling in an undisturbed physiological state.

## Development of the protocol

Recent years have seen the emergence of ingestible devices designed to address this gap. We previously described a capsule-based device (CapScan Gen1) that achieved lumen-targeted sampling in subjects in a normal digestive state^4^. That device consisted of a capsule encasing a folded collection bladder that opened in response to regional pH conditions or timed polymer degradation, allowing capture of luminal contents from a predefined portion of the intestines. These studies demonstrated that SI luminal contents differ substantially from stool and saliva in taxonomic composition, metabolite distribution, bacteriophage induction, and host-derived proteins, while also revealing compositional and functional gradients along the intestinal axis^4,10^.

Despite these advances, first-generation devices had three practical limitations. First, each capsule collected a single luminal sample, requiring multiple ingestions to obtain spatial information. Second, the small devices could be difficult to reliably detect and recover after passage, increasing the risk of device loss burden. Third, because multiple capsules were swallowed independently, the relative ordering between samples could be uncertain within a single individual due to variation in gastric emptying, intestinal transit time, and device activation timing. These factors complicated large-scale, longitudinal, and cohort-based deployment.

To address these challenges, we developed a second-generation device (CapScan^®^ Gen2, Envivo Bio, San Francisco, CA, USA), hereafter referred to as the sampling device. In contrast to Gen1, which collected a single sample at one site, Gen2 functions as a segmented longitudinal collector that captures sequential luminal samples throughout a single physiological transit. The device consists of a 28-cm elastic collection tube (outer diameter, 2.5 mm) attached to a 6-mm head element containing an RFID chip for unique identification and barium sulfate for visibility in X-rays. The tube is packaged within a size 00 hydroxypropyl methylcellulose capsule (22 mm × 8 mm), a size generally swallowable by adults and consistent with regulatory guidance for ingestible devices.

During manufacturing, the collection tube is radially collapsed and wound into multiple concentric layers. Each layer is coated with a distinct enteric coating that dissolves toward the inner layers at either progressively higher pH thresholds or based on time spent in the intestinal tract (**Fig. 1a,b**). As the sampling device traverses the gastrointestinal tract, dissolution of the outermost coating triggers unwinding of that layer. Upon exposure, the elastic tube expands radially and draws in luminal fluid by capillary action. Unwinding of the first layer exposes the second layer of dissolvable polymer to intestinal fluids, priming further sampling, and this process repeats as the device is propelled through the intestines by peristalsis. In practice, up to four visually distinguishable luminal segments are typically recovered, depending on host physiology and transit conditions. The sequentially filled tube stores captured material as a linear array, preserving the relative order in which samples were collected. Gen2 is not designed to assign samples to specific anatomical landmarks (e.g., duodenum, jejunum, ileum). Rather, it enables relative proximal-to-distal sampling during a single physiological transit. Because samples are collected sequentially within the same device, uncertainty in their relative positioning is substantially reduced compared to first-generation approaches. Although small-intestinal motility can include intermittent bidirectional movement, the ordering of captured segments is generally preserved.

**Figure 1:**
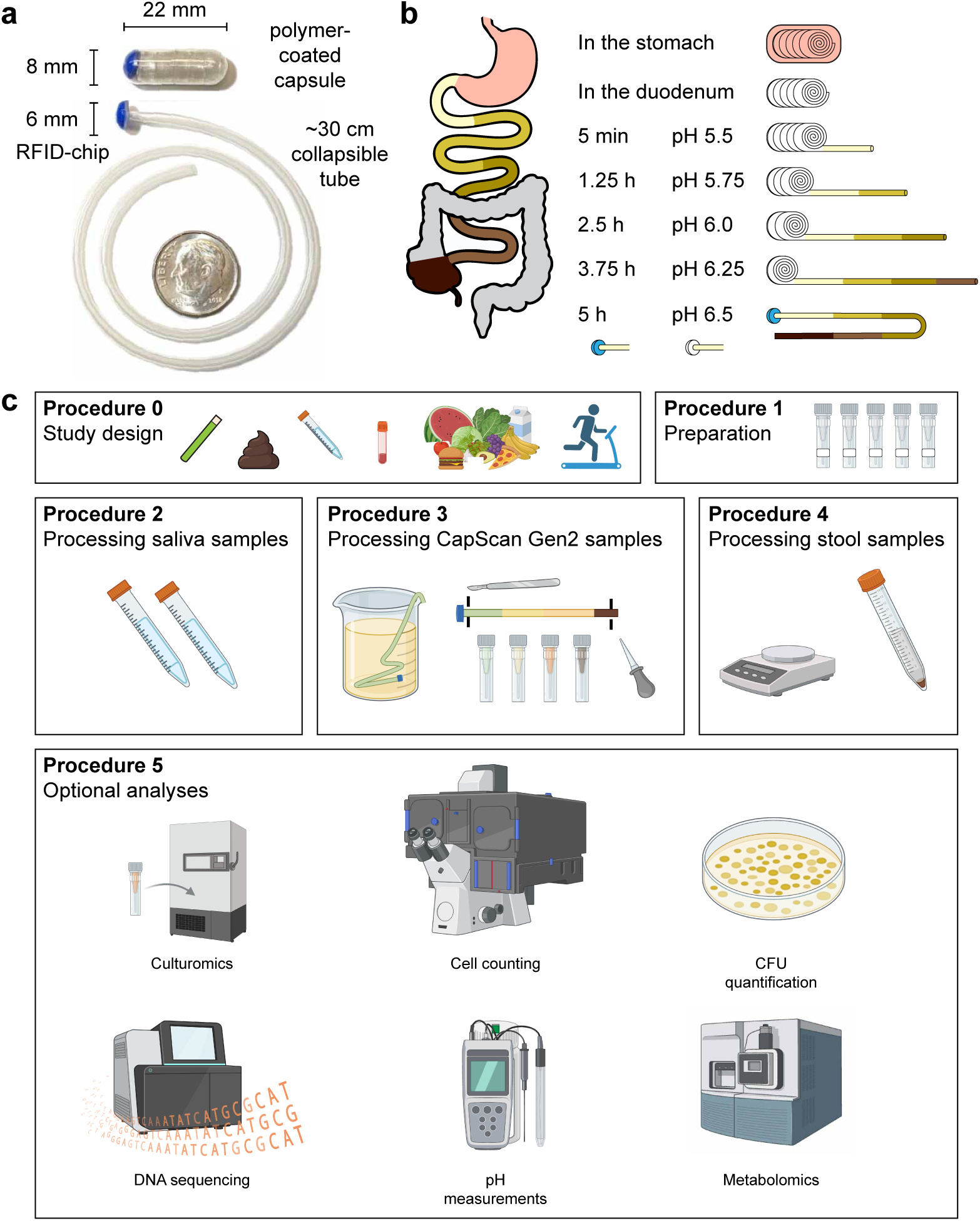
Overview of the CapScan Gen2 device. a) Image showing the polymer-coated capsule encasing a ∼30 cm folded collapsible tube with a blue tip containing an RFID chip. The collection tube is also shown unfolded (U.S. dime shown for scale). b) Schematic illustrating how the tube unfurls and collects sequential luminal samples (∼600 µL) from distinct regions of the GI tract, including the small intestine (SI) and ascending colon. c) Schematics illustrating the major procedures described in this Protocol, including study design, sample preparation, processing of saliva, CapScan Gen2, and stool samples prior to downstream analyses.

**Figure 2:**
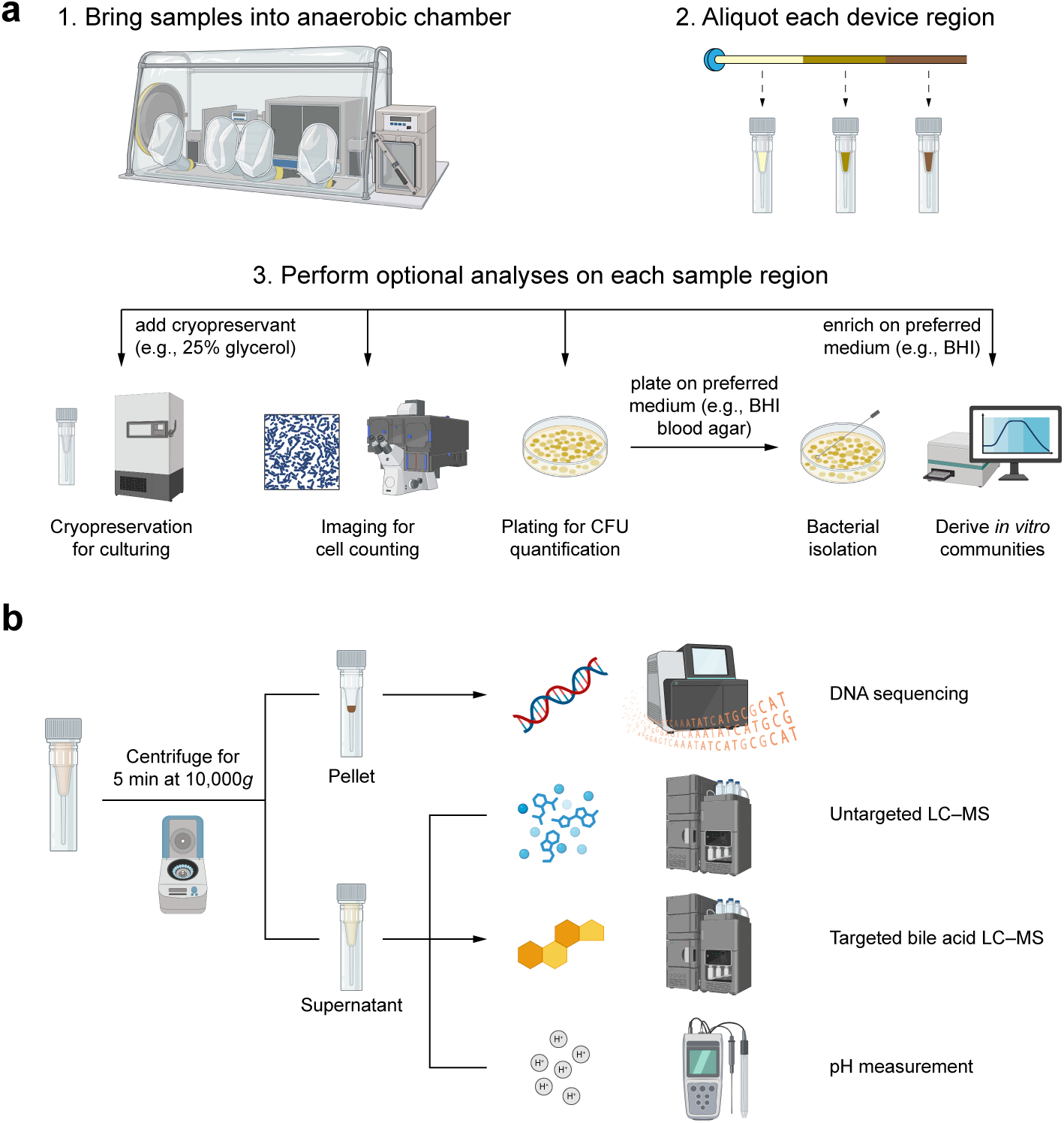
Overview of workflow and downstream analyses enabled by the CapScan Gen2 protocol. a) Workflow for optional anaerobic analyses performed on fresh (non-frozen) device samples. Individual device regions are processed separately inside an anaerobic chamber to preserve microbial viability for cryopreservation, culturing, CFU quantification, bacterial isolation, imaging-based cell counting, and generation of intestine-derived communities. b) Workflow for optional aerobic downstream analyses. pH measurements are ideally performed immediately on raw luminal material prior to centrifugation. Following centrifugation, pellet and supernatant fractions can be allocated to distinct assays, including DNA sequencing/transcriptomics, untargeted LC–MS metabolomics, and bile acid profiling.

Samples are collected and remain in the collection tube via capillary action. Recovered segments are often demarcated by gas pockets; in their absence, the high viscosity of intestinal fluids minimizes mixing and cross contamination between regions. Within-device pH differences and compositional variation are typically preserved across segments, suggesting limited homogenization of contents after capture.

To enable broader spatial coverage, Gen2 devices can be manufactured with distinct activation properties, which differ in the pH threshold of initial activation. Three main designs have been used thus far (indicated by the tip color of the unfurled device): initial sampling in the stomach (red-tipped), proximal SI (white-tipped, pH-sensitive), or distal SI (blue-tipped, time-sensitive). Collection generally ceases once luminal contents become too viscous to enter the device by capillary action, typically within or beyond the ascending colon, and the device will not uncoil further. This Protocol primarily describes workflows for white- and blue-tipped CapScan Gen2 devices. Processing and downstream analyses for red-tipped devices are similar, although these devices begin sampling earlier in the gastrointestinal tract.

As with Gen1, Gen2 devices require no onboard electronics or power source. The extended uncoiled tube facilitates reliable retrieval using a simple hooked capture tool. The embedded RFID chip enables straightforward linkage between recovered devices and ingestion records. A fully filled device yields up to ∼600 µL of luminal fluid (average 335 µL in our studies), distributed across sequential segments. Although per-segment volumes are finite and require prioritization of downstream assays, they are sufficient material for multi-omic analyses when experimental objectives are defined in advance. This Protocol is designed to optimize utilization of the sampled volume.

## Overview of the procedure

In this Protocol, we describe standardized methods for deploying CapScan Gen2 devices in human studies (**Fig. 1c**). We outline study design considerations and handling practices to optimize device performance and sample recovery, procedures for reproducible extraction of luminal contents, and workflows for multi-omic analyses. We include benchmarking assays to quantify microbial cell density (including single-cell imaging-based readouts) and data evaluating intra-device segment variability. Together, these procedures provide a practical framework for implementing ordered SI luminal sampling in human cohort studies.

## Applications

The design of CapScan Gen2 increases throughput, reduces cost, and simplifies use relative to single-region collection devices. Because sequential samples are collected within one device, investigators can examine relative proximal-to-distal variation while reducing logistical complexity associated with repeated capsule ingestions. This facilitates deployment in longitudinal studies, controlled perturbation experiments (e.g., diet, exercise, or drug interventions), and large cohorts under normal digestive conditions to complement existing stool studies.

In early studies, CapScan Gen2 has been used in diverse clinical research contexts, including an intestinal pharmacokinetics study^11^ and an international field-based probiotic intervention study conducted in a cohort of >100 pregnant women (NCT05922683^12^). The ability to align capsule ingestion with dietary, pharmacologic, or behavioral interventions allows investigators to synchronize sampling with defined physiological windows, providing temporal resolution to more directly connect perturbation and response. In addition to microbiome and metabolite measurements, capsule transit time can serve as an informative physiological readout of host gastrointestinal function^13^.

## Comparison to other methods

Sampling the human SI can be achieved through several complementary approaches, including endoscopic, catheter-based, and capsule-based methods. Each method provides distinct advantages and is suited to different experimental questions.

Endoscopic procedures allow direct visualization and targeted sampling of luminal, epithelial, and mucosal-associated compartments^14–17^. Routine esophagogastroduodenoscopy (EGD) with push enteroscopy permits sampling from the duodenum and proximal jejunum, typically up to the ligament of Treitz. Upper double-balloon enteroscopy can extend access further into the distal jejunum and, in some cases, the proximal ileum. Access to the terminal ileum may also be achieved during colonoscopy or lower double-balloon enteroscopy. These approaches enable precise anatomical localization and collection of biopsies and brushings, providing access to host-microbe interfaces that cannot be reached by luminal collectors alone. However, ileal sampling during colonoscopy is performed after bowel preparation and involves retrograde intubation and introduction of gas to expand the lumen, which may alter the native luminal environment or introduce proximal–distal mixing at the time of collection. Moreover, these procedures are invasive, require clinical infrastructure, and are typically performed under fasting conditions, limiting their suitability for repeated, at-home, or diet-responsive sampling under physiological feeding conditions. Repeated balloon enteroscopy may also increase procedural risk, restricting its use in large-scale longitudinal studies or healthy volunteers.

Catheter-based approaches, such as naso-ileal multi-lumen systems, enable dense, location-specific luminal sampling and controlled substrate delivery during feeding interventions^18,19^. These systems provide high temporal resolution at defined sites and are particularly well suited for mechanistic studies of nutrient metabolism^19^. However, they require specialized facilities, are invasive, and are not easily deployed in large cohorts.

A number of ingestible capsule devices have also been developed^20^. Systems incorporating electronics, actuators, or controlled-release mechanisms^21^ enable programmable sampling or release functions, but increased device complexity can raise manufacturing costs and limit scalability. In addition, devices with larger diameters (∼11 mm in some systems^21^) may increase the risk of retention in patients with intestinal strictures, such as those with inflammatory bowel disease. Other passive pH-triggered or magnetically actuated capsules^22–26^ collect discrete luminal samples during transit, typically with modest volumes per activation event. While these systems demonstrate the feasibility of noninvasive GI sampling, challenges can include limited sample yield, reliance on single-region capture per device, and practical issues related to retrieval from stool.

CapScan Gen2 differs conceptually by functioning as a segmented longitudinal collector that captures multiple sequential luminal samples during a single physiological transit. Rather than assigning samples to fixed anatomical landmarks, the device preserves the relative proximal-to-distal order of captured material within one continuous pass through the intestine. This design reduces uncertainty arising from inter-capsule variability in activation timing and simplifies study logistics by enabling ordered multi-segment sampling with a single ingestion. The absence of onboard electronics and the extended uncoiled tube facilitate reliable retrieval and support deployment in longitudinal and cohort-scale studies conducted outside of specialized clinical settings.

Capsule-based luminal sampling is complementary to other measurement modalities. For example, imaging capsules such as PillCam^27^ or gas-sensing capsules such as ATMOS^28^ provide environmental context, while endoscopic biopsies enable tissue-level analysis. Combining these methods with ordered luminal sampling may allow integrated assessment of microbial, chemical, and physiological gradients along the gastrointestinal tract.

## Limitations

Although CapScan Gen2 enables ordered, multi-segment luminal sampling during a single physiological transit, several limitations should be considered when designing studies and interpreting data.

First, sampling occurs during natural gastrointestinal transit and is therefore subject to host-dependent variability. Differences in gastric emptying, SI transit time, hydration status, diet, physical activity, and other physiological factors may influence both the timing and completeness of device activation. Despite protocol modifications such as dietary guidance and hydration control, occasional sampling failures occur. In some cases, the device may activate in the stomach, reducing the number of SI segments collected, although downstream sampling is typically preserved. Incomplete uncoiling or knotting of the collection tube may also occur in a minority of devices, reducing the number of distinguishable segments recovered. In our experience, sampling of at least two distinguishable regions is achieved in >75% of devices, but performance may vary across populations.

Second, the device preserves the relative order of sequentially captured material but does not assign samples to precise anatomical landmarks. Small-intestinal motility includes intermittent bidirectional movement, and retrograde displacement during transit cannot be excluded. In addition, because luminal contents are captured over minutes and retained during transit, microbial growth within the device is possible and may influence biomass measurements or compositional readouts. These factors may blur fine-scale spatial boundaries and should be considered when interpreting quantitative gradients across segments.

Third, CapScan Gen2 primarily captures luminal fluid and suspended material and does not reliably sample mucosal-associated ecosystems/environments or epithelial tissue. As such, it complements rather than replaces endoscopic biopsy-based approaches when mucosal microbiota or host tissue analyses are required.

Fourth, although a fully filled device yields up to ∼600 µL of material, per-segment volumes are finite (typically ≤200 µL). When multiple segments are analyzed to capture spatial gradients, downstream assays must be prioritized according to study objectives.

For example, culturomics, sequencing, metabolomics, and microscopy cannot always be performed simultaneously on every segment. Ingestion of more than one device (designed to either sample the same or distinct regions) may increase spatial coverage and total volume, but at the cost of increased logistical complexity and participant burden.

Finally, as with any human study involving stool handling, subject willingness and volunteer bias may influence recruitment and generalizability. Participants who are comfortable with stool retrieval or motivated by gastrointestinal symptoms may not fully represent the broader population. At the same time, the noninvasive nature of capsule ingestion provides an opportunity to examine how lifestyle variables, including diet, hydration, exercise, stress, and sleep, modulate SI luminal ecology and host characteristics such as transit time under real-world conditions.

Together, these considerations underscore that CapScan Gen2 is best viewed as a tool for sequentially ordered luminal sampling under physiological conditions, and study design should account for these limitations *a priori*. As presently configured, CapScan Gen2 provides a practical platform for investigating SI luminal ecology across space and time in humans. Future device iterations may address current constraints through enhanced location tracking, incorporation of fixation or growth-inhibition strategies^29^, or expanded sampling modalities (e.g., mucus-targeted collection).

## Experimental Design

### Regulatory Approvals

Institutional Review Board (IRB) or equivalent ethics committee approval is required for all studies involving human participants undergoing device administration and sample collection. Protocols should be reviewed to ensure participant safety, informed consent, and compliance with relevant regulations governing investigational medical devices and human biospecimen research.

Material Transfer Agreements (MTAs) should be established when biospecimens or device components are exchanged between collaborating institutions. Investigators should consult institutional regulatory offices early in study planning to determine whether the device is classified as investigational and to clarify institution-specific reporting requirements.

### Study design

Studies employing CapScan Gen2 should be structured to maximize the likelihood of recovering representative luminal samples while minimizing participant burden. Participants should be provided with pre-labeled collection tubes, retrieval tools, stool collection materials, and standardized recording forms (**Fig. 3**) to document capsule ingestion time, bowel movement timing, Bristol stool score, and device recovery time. In longitudinal studies, the RFID serial number of each device should be recorded before ingestion to facilitate linkage of recovered samples to ingestion timing and study interventions. A detailed list of recommended materials is provided in the **Materials** section below. Because gastrointestinal physiology and luminal composition can be influenced by behavioral, dietary, and medical factors, collection of metadata including medical history and habitual diet, as well as monitoring of dietary intake, gastrointestinal symptoms, physical activity, stress, and medication use throughout the sampling period, is strongly recommended to facilitate interpretation of downstream analyses and adjustment for potential confounders.

**Figure 3:**
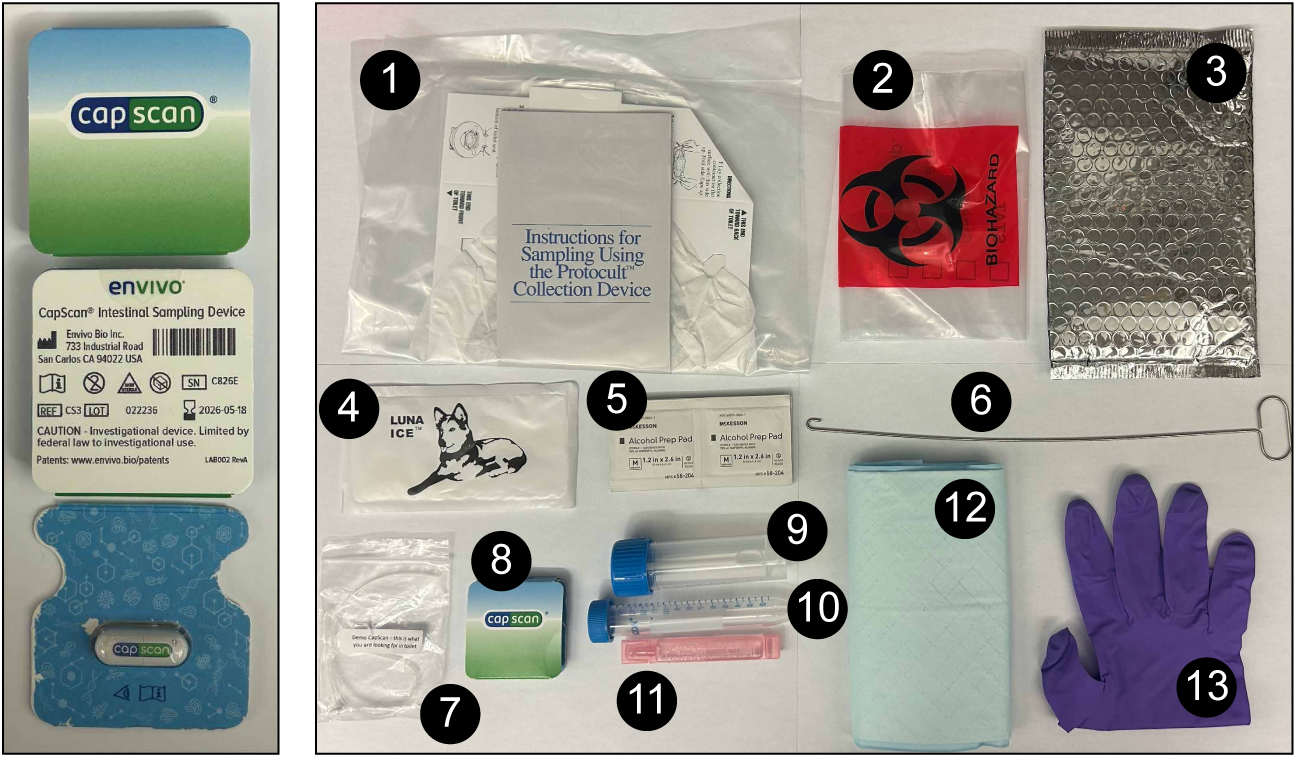
CapScan Gen2 participant collection kit for stool, saliva, and device recovery. The kit contains materials required for at-home sample collection and transport, including a paper-based stool hat (1), biohazard bag (2), thermal transport bag (3), ice pack (4), alcohol prep pads (5), retrieval hook for device recovery from stool (6), demo CapScan device (7), CapScan Gen2 blister pack (8), stool collection tube with spoon (9), saliva collection tube (10), saline solution for saliva collection (11), absorbent pad (12), and disposable gloves (13). Left inset: packaged CapScan Gen2 capsule device within the sealed blister pack.

Evening ingestion is typically recommended, approximately three hours after a meal. Participants consume their usual evening meal and are advised to avoid strenuous exercise and excessive liquid intake for approximately three hours prior to device ingestion. Alcohol consumption should be avoided during the sampling period, as it may alter gastrointestinal transit. A pre-tooth-brushing saliva sample can be collected to provide baseline oral microbiome and metabolite measurements.

The CapScan Gen2 device is swallowed with a glass of water. Participants are encouraged to maintain adequate hydration during the sampling period to increase the likelihood of recovering luminal fluid rather than more distal colonic material. However, drinking too much water before the device transitions into the intestines can delay gastric emptying. After ingestion, participants should inspect each subsequent bowel movement until the unraveled tube is recovered. A disposable metal hook or tweezer may facilitate removal of the tube from stool. When feasible, a paired stool sample should be collected at the time of device recovery.

Unless samples are to be processed immediately (e.g., for culturing or bacterial isolation), devices should be frozen immediately at –20 °C to limit post-recovery microbial growth. Freezing does not disrupt capillary retention of luminal contents within the collection tube, and spatial separation between recovered regions is typically preserved after thawing. Samples can subsequently be transported to the laboratory on ice packs. If fresh samples are required, devices may be temporarily stored at 4 °C and transported with cold packs.

Because device retrieval requires visual inspection of stool, the protocol may be more challenging for participants with frequent bowel movements or loose stools. In settings where access to conventional toilet facilities is limited, alternative collection methods such as stool containers may be required. Stool collection can be facilitated with dissolvable “lily pads” that sit at the water-air interface or paper-based stool hats suspended above the toilet bowl. When dissolvable pads are used, automatic-flush toilets should be avoided to prevent sample loss. Regardless of collection method, care should be taken to avoid contamination of the device and stool sample with urine or toilet water.

SI transit time varies across individuals. In most healthy participants, device passage occurs within 1–3 days, although longer transit times (5–6 days or more) may occur. If capsule passage is delayed, conservative management is generally appropriate, starting with use of a mild laxative. If the capsule has not passed within several days, the device tip contains barium sulfate and is radio-opaque, allowing confirmation of retention by abdominal X-ray if clinically indicated. Participants experiencing symptoms suggestive of obstruction should seek medical evaluation. In our studies, involving thousands of devices, capsule retention requiring intervention has not occurred under defined inclusion and exclusion criteria.

### Participant eligibility

Individuals with known or suspected gastrointestinal obstruction, clinically significant strictures, active diverticulitis, or gastrointestinal malignancy, or prior gastrointestinal surgeries associated with altered luminal anatomy or narrowing should be excluded due to the potential risk of capsule retention. Study-specific inclusion and exclusion criteria should be developed in consultation with clinical investigators and regulatory oversight bodies.

Participants should have no known difficulty swallowing capsules approximately 22 mm × 8 mm in size. Temporary discontinuation of proton pump inhibitors or antacids should be considered for a minimum of one week prior to sampling, as these agents can alter gastric and SI pH and potentially affect device activation; decisions regarding medication management should be made in accordance with clinical guidance and study protocols.

CapScan Gen2 devices have been administered under institutional review in diverse populations, including individuals aged 16–79 years, healthy volunteers, pregnant participants, and patients with gastrointestinal conditions such as inflammatory bowel disease (Crohn’s disease and ulcerative colitis) without known structures, celiac disease, small intestinal bacterial overgrowth, and environmental enteric dysfunction.

### Sample processing for downstream analyses

Analysis of paired saliva and stool samples alongside CapScan devices is helpful for understanding the small intestine in the context of the entire gastrointestinal tract. Saliva and stool samples should be thoroughly homogenized prior to aliquoting. Stool mass should be recorded as wet weight; however, because water content and Bristol stool score vary across individuals, measurement of dry mass may improve normalization across samples.

#### External decontamination and device handling

Careful handling of CapScan Gen2 devices is essential to prevent cross-contamination. Because capsules are excreted in stool, which contains substantially higher microbial density than SI luminal fluid, the external surface of the collection tube represents a potential contamination source. Immediately upon recovery, the open end of the tube should be plugged or clamped to prevent both contamination from fecal material and loss of luminal contents. Excessive bending, squeezing, or compression of the collection tube should be avoided during handling to minimize mixing between adjacent luminal segments. The sealed device can then be submerged in a dilute bleach solution for 15 min to disinfect the exterior surface. Care should be taken to ensure that bleach does not enter the lumen of the tube. After disinfection, the exterior should be cleaned with ethanol wipes prior to sectioning.

#### Content extraction

After exterior cleaning, the tips of both ends are excised with a sterile razor blade. An aspirating bulb attached to a pipette tip is inserted into the distal end of the tube (i.e., opposite the RFID tip), and luminal contents are gently expelled into sterile collection tubes from proximal to distal. This extraction order ensures that lower-density SI contents are expelled first, avoiding contamination with higher-density distal contents. Care should be taken to avoid contact between the tube and the rim of the collection vessels to minimize contamination. Aliquoted samples should be stored in individual tubes rather than shared plates to reduce cross-contamination risk. PCR tubes can be used to conserve freezer space if necessary.

A fully filled device yields approximately 600 µL of luminal material. In practice, droplet counting can provide a rapid estimate of volume: a filled device typically produces ∼24 droplets of ∼25 µL each.

#### Segmentation strategy

Devices can be segmented based on visible changes in color or opacity (a proxy for cell density), the presence of gas pockets (which frequently demarcate discrete sampling events), or by dividing the tube into equal-length sections. Visible demarcations often yield uneven volumes across segments. When uniform fluid volumes are required, equal-length segmentation may be preferable. Alternatively, adjacent segments or multiple devices can be pooled to achieve sufficient volume for downstream analyses, depending on study objectives.

#### Interpreting luminal characteristics

Although CapScan Gen2 preserves the relative order of sequentially captured samples, precise anatomical assignment of individual segments is not possible. Nevertheless, internal content characteristics (color, consistency, and pH) can provide contextual clues. In healthy individuals, gastric contents are highly acidic (pH 1.5–3.5), with pH generally increasing through the SI (duodenum ∼5.8–6.5^30^, ileum ∼7.1–7.9^31^) and decreasing in the cecum (∼6–6.8^31^) before rising again in the colon (∼6.3–7.7)^31–33^. These gradients may vary in individuals with altered acid production (e.g., hypochlorhydria) or other gastrointestinal conditions (e.g., cholestasis), and pH measurements should therefore be interpreted cautiously, particularly in case-control studies involving acid-related pathologies.

Gastric samples are typically liquid, pale yellow-green to pink, and may contain visible fat globules. SI samples vary considerably in appearance but are often dark yellow or orange to light brown and sufficiently fluid to form droplets. Material collected in the ascending colon is usually darker brown and substantially more viscous or sludgy; such samples may not form droplets and may require gentle manual squeezing of the tube for removal. Centrifugation of ascending colon samples often yields minimal supernatant relative to SI samples.

### Maximizing downstream analyses for each segment

CapScan Gen2 samples support a range of downstream analyses, including 16S rRNA gene sequencing, shotgun metagenomics, untargeted metabolomics, targeted mass spectrometry-based analyses (e.g., bile acids, short-chain fatty acids), proteomics, and other host-derived analytes. When devices are maintained unfrozen (e.g., refrigerated shortly after recovery), a substantial fraction of cells will remain viable, enabling culturomics, colony-forming unit (CFU) quantification, microscopy-based cell counting, and downstream *in vivo* studies in animal models.

Because per-segment volumes are finite and device recovery efficiency varies across individuals, investigators should define a prioritization hierarchy before sample processing. An ordered list of analyses aligned with primary study objectives is recommended. **Table 1** provides typical minimum sample volume requirements for common assays based on our experience; these thresholds should be confirmed with individual core facilities.

**Table 1:** Minimum recommended sample volumes for common downstream analyses.

| Assay* | Application | Volume (µL) | Whole sample/homogenate | Clarified supernatant |
| --- | --- | --- | --- | --- |
| A | pH measurement | 5 |  | ✓ |
| B | Culturomics (fresh cultures, isolation, or glycerol stocks) | >20 | ✓ |  |
| C | Microscopy-based cell counting | 1 | ✓ |  |
| D | CFU quantification | 1 | ✓ |  |
| E | DNA sequencing (16S rRNA gene sequencing and/or shotgun metagenomics) | 50<sup>†</sup> | ✓ (or pellet) |  |
| F | Metabolomics | 20–40 |  | ✓ |

Segmentation of the collection tube should be aligned with study objectives. Investigators may partition the device into multiple regions to capture spatial gradients, or alternatively process the entire device as a single bulk SI sample when the primary comparison is between SI and stool (while taking into account potentially large differences in cell density across segments). Even without segmentation, CapScan Gen2 samples consistently differ from paired stool in microbial composition, biomass, and metabolite profiles. However, dividing the device into multiple regions increases spatial resolution and can provide additional biological insight when sufficient volume is available. In our studies, we typically aim to recover three distinguishable regions (R1–R3), each containing ∼200 µL of luminal material. Under these conditions, it is generally feasible to perform culturomics, cell counting, pH measurement, DNA-based sequencing, and metabolomics within a single segment (**Table 1**).

For workflows requiring both cellular and soluble fractions, samples should first be allocated to assays using whole homogenized material (e.g., culturomics, CFU quantification; **Table 1**). Remaining material can then be aliquoted as a fixed volume and centrifuged to separate the pellet and supernatant. The pellet may be used for sequencing instead of homogenized contents if desired (e.g., 16S rRNA gene sequencing or metagenomics), particularly when minimizing host DNA or enriching for cellular biomass is preferred. A visible pellet typically indicates sufficient biomass for DNA-based sequencing. Supernatant allocation should reflect primary study objectives. In more viscous or distal samples, centrifugation may yield a relatively large pellet and minimal supernatant. In such cases, additional purification or extraction steps may be necessary for metabolomics analyses.

When sufficient starting material is available, multiple supernatant-based assays can be performed in parallel from a single centrifuged aliquot by scaling the initial input volume to yield adequate supernatant for all downstream analyses.

### Complementary approaches for estimating absolute microbial cell density

Absolute microbial cell numbers in CapScan Gen2 samples can be estimated using two complementary approaches: single-cell imaging of a defined sample volume and sequencing-based quantification using an exogenous spike-in control. Across a wide range of recovered segments, these approaches typically yielded concordant estimates, differing by only ∼20–30% (**Fig. 4a,b**), which is within expected technical variation for independent absolute quantification methods.

**Figure 4:**
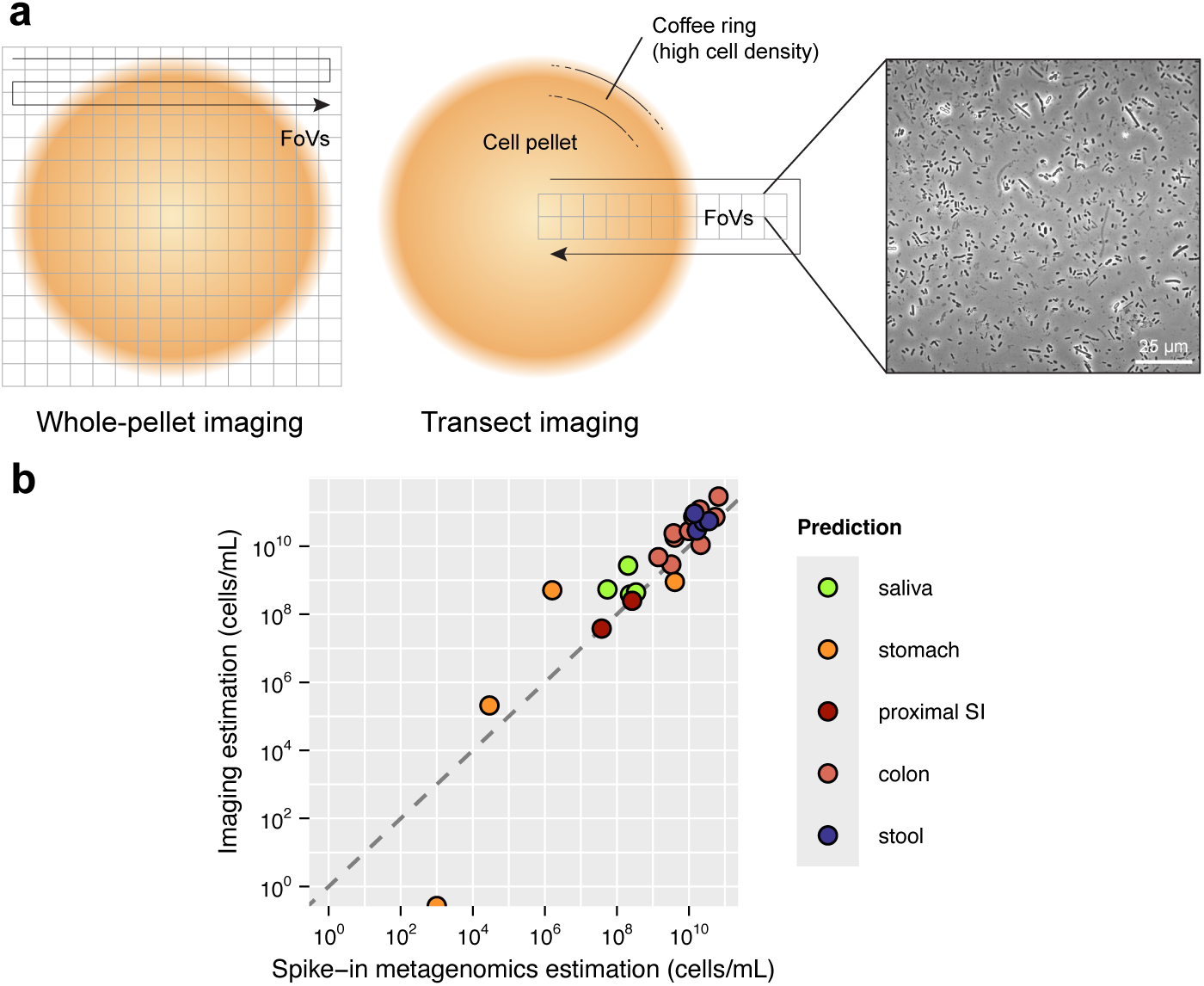
Comparison of imaging-based and sequencing-based estimates of absolute microbial cell density. a) Schematic illustrating imaging-based quantification of microbial cell density from CapScan Gen2 samples. A defined volume of sample (typically ∼1 µL) is spotted onto an agarose pad and imaged across a grid of fields of view (FOVs). A representative phase-contrast image from a single FOV is shown. Microbial abundance can be estimated either by imaging all grid regions or by imaging a transect across the spot, which is faster to acquire and computationally simpler to analyze while still capturing radial variation. Total microbial abundance can then be estimated either by direct cell counting or by quantifying the fraction of image area occupied by cells. Scale bar: 25 µm. b) Comparison of absolute microbial cell density estimates obtained by microscopy-based imaging and sequencing-based spike-in normalization across saliva, stomach, proximal SI, colon, and stool samples. Each point represents an individual sample. Dashed line indicates 1:1 correspondence between the two methods. Across a broad range of microbial biomass values, the two approaches showed close concordance, typically differing by ∼20–30%.

Imaging-based quantification involves spotting a defined aliquot (e.g., 1 µL) of homogenized, diluted sample onto an agarose pad and enumerating fluorescently stained bacterial cells using phase-contrast and epifluorescence microscopy. High-resolution imaging is required to resolve individual bacterial cells; a 100X oil immersion objective is optimal, although a high-numerical-aperture 40X air objective can also suffice. Staining with a DNA-binding dye such as DAPI enables microbial cells to be distinguished from food particles and other debris. Live/dead staining can be used to quantify the fraction of live and dead cells.

Cell enumeration can be performed by imaging the entire circular spot (**Fig. 4a**), which provides the most comprehensive count but requires acquisition of a large number of fields of view (FOVs, typically ∼100–200) and consequently more computational analysis. As a faster alternative, imaging along a single transect (or two orthogonal transects) across the spot can yield accurate estimates with reduced imaging time; a typical transect requires ∼10–25 FOVs depending on camera sensor size (**Fig. 4a**). At minimum, a full radial transect should be captured, as microbial density often varies along the radial axis due to the well-known “coffee ring” effect^34^.

Cell density can then be quantified either by direct enumeration of individual cells or by calculating the area fraction occupied by cells (identified by staining or by phase contrast) within each FOV. The latter provides a relative measure of absolute abundance under the assumption that average cell size does not vary substantially across samples. Numerous open-source and commercial image analysis platforms are available for automated bacterial cell segmentation and counting. When implemented with standardized acquisition parameters, these approaches yield reproducible estimates and can be performed using imaging systems commonly available in microbiology laboratories.

As an orthogonal approach, absolute microbial abundance can be estimated using a defined bacterial spike-in control added prior to DNA extraction and sequencing. The spike-in organism(s) should be taxonomically distinct and not expected to be present endogenously in the sample, ensuring that sequencing reads attributed to the spike-in arise exclusively from the added reference population. This approach, previously applied in stool microbiome studies^35^, enables conversion of relative sequencing reads into absolute abundances.

Because microbial biomass varies substantially across the SI, often spanning several orders of magnitude, spike-in concentrations must be adjusted according to expected sample density (**Table S1**). We typically classify samples as “high-density” or “low-density” prior to extraction based on color, turbidity, consistency, and pH. Samples are considered low density if the color is clear, yellow, turbid yellow, or olive, or if the color is light brown, turbid brown, or turbid olive and pH <5.5. Samples are considered high density if the color is brown and the consistency is sludgy, or if the color is turbid brown and pH >5.5. Incorrect classification can reduce quantification accuracy: excessive spike- in in low-biomass samples may dominate sequencing reads, whereas insufficient spike-in in high-biomass samples may result in inadequate reference counts for precise normalization.

## Materials

Unless otherwise specified, all reagents should be molecular-biology grade. Example suppliers are provided; equivalent products from other manufacturers may be substituted.

### Collection kit provided to participants

- Brown paper bags (e.g., Reli. Store, cat. #B09KYBZQRT)
- Biohazard specimen bags (e.g., keebomed, cat. #B0CV8X477H)
- Absorbent pads (e.g., Stack Man Store, cat. #B0DX4WDBX9)
- 15-mL conical tube for saliva collection (e.g., Corning, cat. #352196)
- Stool collection tube (e.g., Globe Scientific, cat. #109120)
- 5-mL sterile saline ampule (e.g., Bangerz Sunz, cat. #B08NQ38KQ1)
- Insulated transport bag (e.g., Amazon Basics Store, cat. #B093WNNDLD)
- Reusable ice pack (e.g., Luna Ice, cat. #B07WS3JPFP)
- Nitrile gloves (e.g., Med Pride, cat. #B00GS8VGP6)
- Alcohol wipes (e.g., PhysiciansCare, cat. #51019-001)
- Metal retrieval hook or tweezers (e.g., SE Store, cat. #365PT12)
- Aluminum insulated bag (e.g., Smilco Store, cat. #B08JHS66VG)
- Stool collection hats or dissolvable pads (e.g., Fisher Scientific, cat. #50-465-371)
- Styrofoam transport box (e.g., Fisher Scientific, cat. #NC1494675)
- CapScan Gen2 device
- Printed instructions

### Equipment

#### Sample processing (Procedures 1–4)

- High-precision milligram digital scale (e.g., Smart Weigh, cat. #SW-GEM50)
- Vortex mixer (e.g., Vortex-Genie 2, Scientific Industries, cat. #SI-0236)
- RFID reader (e.g., GIGA-TMS Inc, cat. #MP101A)
- Laptop or equivalent device for recording sample metadata (**Table S2**)
- Camera for documenting samples (smartphone camera is acceptable)
- Anaerobic chamber (e.g., Coy Laboratories Type A Vinyl Anaerobic Chamber, cat. #7000040R)
- Microcentrifuge (e.g., ThermoFisher, cat. #75002436)

#### Downstream analyses (Procedure 5)

- Anaerobic chamber (e.g., Coy Laboratories Type A Vinyl Anaerobic Chamber, cat. #7000040R) *[also used in processing]*
- Microcentrifuge (e.g., ThermoFisher, cat. #75002436) *[also used in processing]*
- Micro-pH sensor (e.g., Ultra-micro-ISM, Mettler Toledo, cat. #30244732, and Micro-Combination pH Probe (2 cm), Microelectrodes, cat. #MI-414E)
- Anaerobic chamber (e.g., Coy Laboratories Type A Vinyl Anaerobic Chamber, cat. #7000040R)
- Microcentrifuge (e.g., ThermoFisher, cat. #75002436)
- Phase-contrast microscope (e.g., Ti-E, Nikon Instruments) with a high NA objective (≥40X) and a camera (e.g., Neo 5.5 sCMOS camera, Andor Technology) for single-cell imaging

### Reagents

#### Chemicals

##### Sample processing (Procedures 1–4)

- 70% ethanol (EtOH; Fisher Scientific, cat. #BP82011)
- Sodium hypochlorite (bleach) solution (Essendant, cat. #CLO30966CT)

##### Downstream analyses (Procedure 5)

- 70% ethanol (EtOH; Fisher Scientific, cat. #BP82011) *[also used in processing]*
- Sodium hypochlorite (bleach) solution (Essendant, cat. #CLO30966CT) *[also used in processing]*
- Bovine serum albumin (BSA; VWR, cat. #0332-25G)
- Tween 20 (Promega, cat. #H5151)
- LIVE/DEAD™ *Bac*Light™ Bacterial Viability kit (Invitrogen, cat. #L13152)

#### Media and buffers

##### Sample processing (Procedures 1–4)

- Sterile 50% (v/v) glycerol solution (Teknova, cat. #G1796)
- 1X phosphate-buffered saline (PBS), sterile-filtered (Fisher Scientific, cat. #J61196-AP)

##### Downstream analyses (Procedure 5)

- 1X phosphate-buffered saline (PBS), sterile-filtered (Fisher Scientific, cat. #J61196-AP) *[also used in processing]*
- Sterile 50% (v/v) glycerol solution (Teknova, cat. #G1796) *[also used in processing]*
- Difco Brain Heart Infusion (BHI) medium (Becton Dickinson, cat. #237200)
- Difco granulated agar (Becton Dickinson, cat. #214510)
- Defibrinated horse blood (Hardy Diagnostics, cat. #SHS500)
- Nuclease-free water (Fisher Scientific, cat. #AM9932)

#### Plasticware and consumables

- 1000-µL LTS-filtered pipette tips (Rainin, cat. #30389212)
- 200-µL LTS-filtered pipette tips (Rainin, cat. #30389239)
- 20-µL LTS-filtered pipette tips (Rainin, cat. #30389225)
- 200-µL LTS-filtered pipette tips, wide-orifice 1.5 mm (Rainin, cat. #30389241)
- Biotix uTIP P200 filter tips (Fisher Scientific, cat. #12-111-126)
- Aspirating bulbs (Yohii, cat. #B07FM8B8FK)
- Absorbent underpads (Fisher Scientific, cat. #14-206-62)
- Disposable sterile scalpel blades (Med Pride, cat. #UPS 352410471018, or Slice, cat. #10524)
- 1-mL screw-cap microcentrifuge tubes with O-ring (VWR, cat. #10025-748)
- 500-µL screw-cap microcentrifuge tubes with O-ring (VWR, cat. #10026-098)
- 15-mL centrifuge tubes, conical bottom (Corning, cat. #352196)
- 10-mL sterile serological pipettes (Celltreat, cat. #229210B)
- PCR strip tubes (ThermoFisher, cat. #AB-0264)
- Reservoirs for liquid handling (ThermoFisher, cat. #9510037)
- Cryo-Babies CRYOtags for laser printer (Diversified Biotech, cat. #LCRY-1700)
- Tough Spots tube labels (Fisher Scientific, cat. #50-550-261)
- Hydrion pH indicator strips (MicroEssential, cat. #67)
- 70% isopropanol wipes (First Aid Only, cat. # 51019-001)
- 100 mm × 15 mm polystyrene Petri dishes (Fisher Scientific, cat. #FB0875713)
- Cryogenic/freezer storage boxes (Fisher Scientific, cat. #03-395-465)
- 50-mL conical tubes (Corning, cat. #352070)
- Mini binder clips (Fisher Scientific, cat. #50-205-1951)
- Inoculating loops (Fisher Scientific, cat. #22-363-597)
- Kimwipe task wipers (Fisher Scientific, cat. #06-666)
- Bleach wipes (PDI, cat. #U26595)
- Flanged nylon plugs compatible with tubing diameter (EldonJames, cat. #P0-1-200BN)

### Equipment setup

#### Preparation of anaerobic chamber

If preservation of viable anaerobic microbes is desired (e.g., for culturomics, CFU quantification, or live-cell imaging; **Fig. 2**), BHI plates supplemented with 1.5% (w/v) agar and 5% blood, liquid media, and all required plasticware should be transferred into the anaerobic chamber at least 48 h prior to use. To permit gas exchange, loosen caps on liquid containers to permit gas exchange during this period. Allow plastics and media to equilibrate within the chamber for 48–72 h before use. CRITICAL All materials introduced into the anaerobic chamber should be allowed to equilibrate for a minimum of 48 h to ensure adequate oxygen removal. Introducing materials immediately before use may result in residual O_2_ exposure and reduced viability of strict anaerobes. To minimize O_2_ introduction into the chamber, plastics and liquids can be transferred while still warm following autoclaving. If viable cell preservation is not required, sample processing can instead be performed aerobically in a biosafety cabinet.

The following materials should be equilibrated inside the anaerobic chamber:

- 500-µL and 1-mL screw-cap microcentrifuge tubes with O-ring
- 10-mL sterile serological pipettes
- P1000, P200, and P20 LTS filter-tip boxes
- Wide-orifice P200 LTS filter-tip box (for viscous samples)
- Biotix uTIP P200 filter tips compatible with the aspirating bulb
- Absorbent pads
- Disposable sterile scalpel blades
- 15-mL conical centrifuge tubes
- Tube racks
- 100 mL of 1X PBS
- 50 mL of 50% glycerol
- 70% isopropanol wipes
- Analytical balance
- Tweezers

### Procedure

Approximate timing estimates are provided for processing a single CapScan Gen2 device and its associated saliva and stool samples. When multiple devices are processed, many steps can be performed in parallel to improve workflow efficiency and reduce overall time per device.

To minimize cross-contamination between high- and low-biomass sample types, we recommend processing saliva, stool, and CapScan samples separately rather than simultaneously. In particular, CapScan samples are ideally processed in a dedicated low-biomass biosafety cabinet or anaerobic chamber, physically separated from areas used for stool aliquoting and processing. Processing different sample types on separate days further reduces the risk of aerosol or surface-based contamination.

### Procedure 1: Preparing for sample aliquoting and processing

TIMING ∼30 min

1. Remove any frozen samples of the selected type to be processed that day (e.g., saliva, CapScan devices, or stool samples) from –20 °C or –80 °C storage and thaw at room temperature until fully liquefied. Once thawed, keep all samples on ice during processing.

2. Print CryoBaby labels in advance. A single CapScan Gen2 device typically yields sufficient volume for 2–3 regions; we recommend planning for three regions (R1–R3). Each region will be aliquoted into multiple tubes for downstream analyses. The example below includes five tubes per region; additional tubes may be required depending on planned assays.

i. Labels ideally should include: sample ID, subject/kit ID, collection date, sample type (i.e., saliva, CapScan, stool), region (for CapScan samples only), and analysis type (e.g., sequencing: SEQ, excess sample: EX, metabolomics: MB, bile acid analysis: BA, glycerol stock: GS), where colors match those of the round CryoBaby labels in Step 4).

ii. Example labels:

- LS0367_Subj1_240906_saliva_SEQ
- LS0457_Subj2_240823_capscan_R3_MB

In this example, LS0457 denotes the sample ID, Subj2 the subject ID, 240823 the collection date, CapScan the sample type, R3 the region, and MB the intended analysis type.

3. Apply rectangular CryoBaby labels to the lower half of each 500-µL screw-cap tube with O-ring, ensuring that sample contents remain visible after filling.

4. Label round CryoBaby labels manually with the sample ID and apply them to the tube cap. Match label colors to the designated analysis type to facilitate rapid identification during processing.

5. Pre-label binder clips with the subject/kit ID and collection date. These will be used to clamp devices closed during processing. Flanged plugs may be used as an alternative, although they cannot be labeled directly.

6. Pre-label microcentrifuge tubes designated for storage of the RFID-containing capsule tip with the subject/kit ID and collection date.

### Procedure 2: Processing saliva samples (if collected)

TIMING ∼5 min

7. Thoroughly homogenize the saliva sample by pipetting up and down. Due to sample viscosity, wide-bore P200 tips may be required.

8. Aliquot appropriate volumes into pre-labeled 500-µL screw-cap tubes with O-rings according to planned downstream analyses.

9. Store aliquots at –80 °C in a freezer box.

10. Update the electronic sample metadata log.

### Procedure 3: Processing CapScan Gen2 samples

TIMING ∼30 min

#### Cleaning devices

TIMING ∼20 min

CRITICAL Perform the following steps in a fume hood or biosafety cabinet due to adhered stool. These steps are typically conducted aerobically but may also be performed inside an anaerobic chamber if required.

11. Decontaminate hood surfaces with 70% ethanol. Cover the work surface with a blue absorbent pad. Prepare a biohazard waste container lined with a biohazard bag.

12. Prepare ∼200 mL of fresh 10% bleach in a glass beaker, sufficient to submerge the device. Square plastic containers (e.g., disposable food-storage containers) can also be used and may facilitate simultaneous processing of multiple devices by providing greater surface area. CRITICAL Bleach degrades over time, so prepare fresh solution (≤1 week old).

13. Remove the capsule device from its collection bag and place it on the absorbent pad inside the hood.

14. Clamp the open end of the device shut using a pre-labeled binder clip. Gently wipe the tube near the opening with a bleach-soaked Kimwipe to avoid clamping residual stool, being careful not to disturb the internal regions. Alternatively, insert a sterile flanged plug to seal the tube.

15. While holding the sealed end upward, submerge the device in 10% bleach for 15 min. CRITICAL Do not allow the clamped (open) end to contact with the bleach unless using a fully sealed flanged plug.

16. Remove the device and wipe it thoroughly with a Kimwipe sprayed with 70% ethanol until all visible stool is removed.

17. Transfer the cleaned device to a fresh absorbent pad and loosely wrap it until ready for sectioning. If processing multiple devices simultaneously, place cleaned devices on ice until processing.

#### Segmenting devices into distinct regions

TIMING ∼10 min

CRITICAL If the device was previously frozen, skip step 1 and perform all subsequent steps aerobically in a biosafety cabinet. Optional viability-based assays (glycerol stocks, culturing, CFU quantification, and single-cell imaging of live cells) should not be performed since freezing may render cells nonviable.

18. For fresh samples intended for viability-based analyses, transfer the cleaned device into the anaerobic chamber using a low-vacuum protocol (e.g., vacuum limit 6 inHg, 9 purge cycles, 9 gas-mix cycles). CRITICAL Low vacuum prevents rapid depressurization and expulsion of contents.

19. Place each device on an individual sheet of white paper labeled with subject/kit ID and collection date.

20. Re-clean the device exterior with ethanol wipes, paying particular attention to the region near the tip to prevent contamination with stool contents during sectioning.

21. Plan incision sites. Label regions sequentially from the tip (R1, R2, R3, …, X = discard).

a. Regions may be defined by visible gas pockets, abrupt color changes, differences in opacity (a proxy for cell density), or other apparent transitions.

b. Regions may also be combined to achieve volumes >200 µL to support prioritized downstream analyses.

22. Photograph the intact device prior to cutting to document visible internal features (e.g., gas bubbles, color gradients, and segment boundaries) for future reference.

23. Using a sterile scalpel, cut close to the RFID-containing tip. Place the excised tip into the pre-labeled microcentrifuge tube.

24. Wipe the scalpel with an alcohol wipe before the next cut.

25. Make a second cut at the distal end of the tube to remove the terminal region (X), which typically contains ascending colonic material and is discarded unless otherwise specified.

26. Attach a sterile Biotix uTIP P200 filter tip to a hand vacuum bulb. Insert the tip into the distal opening of the tube.

27. Gently expel the contents of each region sequentially (typically ∼200 µL per region) into the corresponding EX-labeled 500-µL screw-cap tubes with O-rings. CRITICAL Ensure the capsule tubing does not contact the receiving tube to prevent exterior contamination. This precaution is particularly important given the large difference in microbial density between SI samples (10^3^–10^8^ cells/mL) and stool (10^11^–10^12^ cells/mL).

a. Record the number of droplets expelled to estimate region volume (typically ∼8–10 drops per region).

28. Photograph each EX-labeled tube, ensuring that the sample ID is visible.

29. Aliquot appropriate volumes into analysis-specific tubes according to study priorities (see **Procedure 5** below).

30. Store all aliquots in a freezer box at –80 °C.

31. Scan the RFID chip and record the serial number in the metadata log.

32. Update the electronic metadata log and upload photographs to a secure, PHI-compliant storage location.

### Procedure 4: Processing stool samples

TIMING ∼20 min

CRITICAL If the stool sample was previously frozen, skip step 1 and perform all subsequent steps aerobically in a biosafety cabinet. Optional viability-based analyses (glycerol stocks, culturing, CFU quantification, and single-cell imaging) should not be performed on previously frozen samples since freezing substantially reduces cell viability.

33. For fresh samples intended for viability-based analyses, transfer the stool tube into the anaerobic chamber.

34. Pre-tare a 15-mL conical tube and record the weight.

35. Homogenize the stool thoroughly with a sterile serological pipette.

36. Transfer ∼0.5 g of stool into the pre-tared 15-mL tube and record the mass.

37. Resuspend stool 1:10 (w/v) in sterile, anoxic 1X PBS (e.g., 1 g of stool in 10 mL of PBS). Vortex vigorously for 30 s.

38. Allow large particulates to settle for 5 min.

39. Transfer 200 ± 20 mg of homogenized stool into EX-labeled 500-µL screw-cap tubes with O-rings.

40. Aliquot appropriate volumes or masses into analysis-specific tubes according to study priorities.

41. Store aliquots in a freezer box at –80 °C.

42. Store remaining original and resuspended stool in sealed biohazard bags at –80 °C.

43. Update the electronic sample metadata log.

### Procedure 5: Optional analyses

#### A: pH measurements (CapScan, stool)

TIMING ∼1 min per sample

CRITICAL To obtain physiologically relevant measurements, pH should be measured immediately after sample extraction and prior to centrifugation or supernatant isolation. Delayed measurements or measurements performed on clarified supernatants may yield artificially elevated pH values due to off-gassing, continued microbial metabolism, and loss of mucus-associated buffering capacity.

44. For saliva samples:

a. Measure pH directly in homogenized saliva using either pH strips or a calibrated micro-pH probe. Record the value in the metadata log

45. For CapScan samples:

a. Measure pH directly in the raw homogenized luminal sample using either pH strips or a calibrated micro-pH probe. Record the value in the metadata log.

46. For stool samples:

a. Transfer 150±15 mg into a temporary microcentrifuge tube.
b. Insert the pH probe directly into the sample and record the value.

47. Probe cleaning and decontamination:

a. After each measurement, immerse the probe in 10% bleach to loosen adhered material.
b. Gently wipe the probe clean with a Kimwipe.
c. Rinse the probe by immersing it in 50 mL of Milli-Q water for approximately 5 s.
d. Gently wick away residual liquid from the probe tip using a Kimwipe before proceeding to the next sample.

#### B: Culturomics and microbial community banking (fresh CapScan or stool samples)

TIMING ∼15 min (for setup only)

CRITICAL Perform these steps in an anaerobic chamber to preserve cell viability.

48. Prepare glycerol stocks for long-term storage:

a. For CapScan samples, transfer 20 µL of homogenized sample into a GS-labeled 500-µL screw-cap tube with O-ring containing 20 µL of sterile 50% (v/v) glycerol. Mix gently.
b. For stool samples, transfer 500 µL of the 1:10 PBS-resuspended stool into a GS-labeled 1-mL tube containing 500 µL of sterile 50% (v/v) glycerol. Mix gently.

49. Store glycerol stocks at –80 °C.

50. To derive stable *in vitro* communities (optional), inoculate 1–10 µL of sample into 200 µL of anaerobic growth medium (typically BHI supplemented with 500 mg L^−1^ L-cysteine, 5 mg L^−1^ vitamin K3, and 5 mg L^−1^ hemin) and incubate at 37 °C for 48 h. See ref. ^36^ for detailed community stabilization protocols.

#### C: Cell density estimation by microscopy (fresh CapScan or stool samples)

TIMING ∼60 min

CRITICAL Perform these steps in an anaerobic chamber to preserve cell viability.

51. Because microbial densities vary substantially between SI and stool samples, prepare multiple serial dilutions to ensure accurate imaging-based quantification.

a. Transfer 100 µL of each CapScan region or 1:10 stool PBS resuspension into the top row of a sterile 96-well PCR plate.

i. The undiluted sample well serves as a temporary source for imaging-based quantification. After aliquots are removed for dilution and imaging, the remaining undiluted material should be transferred to SEQ-labeled 500-µL screw-cap tubes with O-rings for downstream sequencing analyses.
ii. The 1:10 stool PBS resuspension is used for cell counting and CFU quantification but not directly for sequencing.
b. Prepare a 1:100 dilution by transferring 1 µL of sample into 99 µL of PBS. Mix thoroughly.
c. Prepare a 1:1,000 dilution by transferring 10 µL of the 1:100 dilution into 90 µL of PBS. Mix thoroughly.
d. For technical duplicates, transfer 50 µL from each dilution well into a corresponding well in the lower half of the plate (well A1 to well E1, well B1 to well F1, etc.).

52. Acquire phase-contrast and Live/Dead fluorescence images.

a. Prepare a 1% (w/v) PBS agarose pad containing Live/Dead stain.

i. Dissolve agarose (1% (w/v)) in PBS and melt by microwaving.
ii. Before the agarose solidifies, add SYTO 9 and propidium iodide to final concentrations of 0.6 µM and 3 µM, respectively, and cast an agarose pad.
b. Gently spot 1 µL of each diluted sample onto the agarose pad. Allow the droplet to dry until cells settle into a roughly circular spot, then place a coverslip on the pad.
c. Acquire phase-contrast and fluorescence images for each sample. Define the imaging grid to cover the entire spot, or alternatively, image along several transects of the spot to account for radial cell density variation caused by the coffee ring effect. Full-spot imaging provides the most accurate estimates but substantially increases acquisition and analysis time.

53. Segment cells in phase-contrast images and calculate cell density.

a. Segment phase-contrast images using DeepCell^37^, a machine-learning-based segmentation tool trained on annotated images of CapScan and stool samples.
b. Compute cell density either from direct cell counts or from the estimated fraction of each field of view occupied by cells.
c. Quantify the fraction of live and dead cells using the Live/Dead fluorescence images and the cell contours obtained from DeepCell segmentation.

#### D: Colony forming unit (CFU) quantification (fresh CapScan or stool samples)

TIMING ∼15 min (excluding incubation and post-incubation steps)

CRITICAL Perform all steps in an anaerobic chamber to preserve cell viability.

54. Because microbial densities differ by several orders of magnitude between SI and stool samples, prepare serial dilutions appropriate for each sample type.

55. For CapScan samples, plate 1 µL of each CapScan region directly onto BHI–5% blood agar plates using an L-spreader.

a. If expected density is ∼10^4^ CFU mL^−1^, plating 1 µL yields ∼10 colonies.
b. If expected density exceeds 10^5^ CFU/mL (>100 colonies per µL plated), prepare additional serial dilutions (e.g., 1:10, 1:100, 1:1,000).

56. For stool samples, starting from the 1:10 PBS resuspension:

a. Prepare a 1:10,000 dilution by transferring 1 µL of the 1:10 resuspension into 1 mL of PBS.
b. Prepare a 1:10,000,000 dilution by transferring 1 µL of the 1:10,000 dilution into 1 mL of PBS.
c. Plate 1 µL of the 1:10,000,000 dilution onto BHI–5% blood agar using an L-spreader.

i. If stool density is ∼10^12^ CFU mL^−1^, this yields ∼100 colonies.
ii. If expected density is <10^11^ CFU mL^-1^ (<10 colonies plated), use a lower dilution (e.g., 1:10,000, 1:100,000, 1:1,000,000).

57. Incubate plates anaerobically at 37 °C overnight.

58. Photograph plates at 24 h. For slow-growing organisms, continue incubation for up to 72 h, photographing every 24 h.

59. Count colonies using the image with the highest number of clearly distinguishable, non-overlapping colonies.

#### E: DNA sequencing sample allocation (Saliva, CapScan, stool)

TIMING ∼10 min (sample allocation only; extraction not included)

60. For saliva samples:

a. Transfer 200 µL of homogenized saliva into SEQ-labeled 500-µL screw-cap tubes with O-rings. Wide-bore P200 tips may be required due to viscosity.

61. For CapScan samples (Option 1: Pellet):

a. Transfer 100 µL from each region (R1–R3) in the EX-labeled tubes into the corresponding SEQ-labeled tubes.
b. If sample volume is limited or material is visibly particulate, centrifuge SEQ-labeled tubes at 10,000*g* for 5 min.

i. The pellet can be used directly for DNA extraction (16S rRNA gene sequencing and/or metagenomics).
ii. The supernatant can be reserved for metabolomics, host-analyte profiling, or other downstream analyses.

62. For CapScan samples (Option 2: Full sample):

a. Transfer 50 µL from each region (R1–R3) in the EX-labeled tubes into the corresponding SEQ-labeled tubes.

63. For stool samples:

a. Pre-tare a SEQ-labeled 500-µL screw-cap tube with O-ring.
b. Transfer 150 ± 15 mg of stool into the tube using a sterile scoop and a pipette tip. Record the mass.

64. DNA extraction should be performed using a validated protocol appropriate for low-biomass (CapScan) and high-biomass (stool) samples. Both 16S rRNA gene sequencing and shotgun metagenomics are compatible with material collected using this protocol. Investigators should follow manufacturer recommendations and include negative controls to monitor potential contamination, particularly for low-biomass SI samples.

65. Optional – If using an extrinsic spike-in to quantify absolute microbial cell density.

a. Prepare diluted ZymoBIOMICS™ Spike-in Control I with molecular-grade water to the appropriate sample type as shown in **Table S1**. Add the spike-in to the sample. Proceed with DNA extraction according to the manufacturer’s protocol.

#### F: Metabolomics (saliva, CapScan, stool)

TIMING ∼10 min

66. For saliva samples:

a. Transfer 50 µL of homogenized saliva into the appropriate 500-µL screw-cap tube with O-ring (e.g., MB for metabolomics, BA for bile acid profiling). Wide-bore P200 tips may be required due to viscosity.

67. For CapScan samples:

a. Transfer 50 µL from each EX-labeled region tube into a temporary microcentrifuge tube.
b. Centrifuge at 10,000*g* for 5 min to pellet particulate material.
c. Transfer 40 µL of clarified supernatant into the appropriate metabolomics analysis tube (e.g., MB or BA)

68. For stool samples:

a. Pre-tare the appropriate labeled tube.
b. Transfer 100±10 mg for untargeted metabolomics (MB) or 200±20 mg for bile acid analysis (BA) using a sterile scoop and pipette tip.
c. Record the mass.

69. Metabolomics samples should be processed using a validated extraction protocol appropriate for aqueous luminal fluid or stool matrices. Typical workflows include protein precipitation using a cold organic solvent (e.g., methanol or acetonitrile), centrifugation to remove debris, and transfer of supernatant for LC–MS analysis. Internal standards and pooled QC samples should be included to monitor extraction efficiency and instrument drift. Investigators should validate extraction methods for low-volume SI samples prior to large-scale studies.

## Timing

Procedure 1 (Steps 1–6), preparation for sample processing: ∼30 min

Procedure 2 (Steps 7–10), saliva processing: ∼5 min

Procedure 3 (Steps 11–32), CapScan Gen2 processing: ∼30 min

Cleaning devices (Steps 11–17): ∼20 min

Segmenting into regions (Steps 18–32): ∼10 min

Procedure 4 (Steps 33–43), stool processing: ∼20 min

Procedure 5, optional analyses:

A. pH measurements (Steps 44–45): ∼1 min per sample
B. Culturomics (Steps 48–50): ∼15 min (excluding culture incubation)
C. Cell counting by microscopy (Steps 51–53): ∼60 min (excluding image analysis)
D. CFU quantification (plating only, Steps 54–59): ∼15 min (excluding incubation; incubation 24–72 h)
E. DNA sequencing sample allocation (Steps 60–65): ∼10 min (extraction not included)
F. Metabolomics sample allocation (Steps 66–69): ∼10 min (extraction not included)

### Troubleshooting

Troubleshooting guidance is summarized in **Table 2**.

**Table 2:** Troubleshooting guide for CapScan Gen2 device recovery and sample processing.

| Step | Problem | Possible reason | Solution |
| --- | --- | --- | --- |
| 13 | Device tubing is broken or leaking | Mechanical damage during recovery or processing | Take a photograph of the damaged region. Cut around the break and clamp the tubing shut. Use bleach wipes instead of submerging the device in bleach solution to avoid bleach entering the tubing lumen. |
| 13 | Device tubing appears knotted | Rare coiling or tangling during intestinal transit | Carefully separate and unknot the tubing using tweezers, taking care not to puncture or compress the collection tube. |
| 16 | Stool adhered to CapScan Gen2 device | Low Bristol stool score (hard, dry stool) | Gently wrap the device in Kimwipes soaked in 70% ethanol and allow it to sit for several minutes before carefully removing adherent material. Avoid excessive mechanical force. |
| 21 | Low sample volume recovered from the device | Incomplete uncoiling or knotting of the device | Process the entire device as a single bulk SI sample instead of segmenting into multiple regions. |
| 27 | Device contents are highly viscous or sludgy and do not readily form droplets | Sample collected in the ascending colon | Gently squeeze the tubing manually to facilitate sample removal. |
| 29 | Device contents cannot be pipetted efficiently | Sample contains food-derived particles or other debris | Use a wide-bore pipette tip. |
| 61b | No visible pellet after centrifugation | Low microbial biomass in proximal SI or stomach-derived samples | Increase input volume for downstream assays or concentrate the sample where possible. Confirm biomass using imaging-based quantification if needed. |

### Anticipated results

#### Reproducible quantification of microbial cell density

Absolute microbial cell densities estimated by single-cell imaging and sequencing spike-in normalization typically show close agreement, differing by ∼20–30% across CapScan Gen2 samples (**Fig. 4a**). This level of concordance falls within expected technical variability for independent quantification methods and supports the use of either approach depending on study design and available resources.

Within individual devices, microbial cell density often varies substantially across recovered segments, frequently spanning multiple orders of magnitude from proximal to distal regions. Such gradients are expected and reflect spatial heterogeneity along the small intestine. Uniform cell densities across all segments are uncommon and, when observed, may indicate inaccurate identification of region boundaries during device processing (i.e., cutting at incorrect transition points between regions) and/or sample mixing.

When compared to paired stool samples, SI segments generally exhibit markedly lower biomass, particularly in proximal regions. Visible pellet formation after centrifugation typically correlates with higher cell density and successful downstream sequencing. Together, these patterns provide internal consistency checks for device performance and segmentation quality.

#### Benchmarking spatial variation along a device

To assess whether standard three-region segmentation (R1–R3) captures the majority of spatial heterogeneity within a CapScan Gen2 device, we performed higher-resolution sectioning in which devices were subdivided into multiple smaller sequential segments, following a similar procedure to that described in steps 21–28.

Across these benchmarking experiments involving devices sectioned into 7-8 regions, microbial composition, CFU counts, pH measurements, and microscopy-based cell density measurements revealed pronounced spatial structure along individual devices. In both subjects, more distal regions were darker in appearance (**Fig. 5a,b**), exhibited modestly higher pH values (**Fig. 5c**), and displayed microbial community compositions that more closely resembled paired stool samples (**Fig. 5d,e**), consistent with sampling of the ascending colon. CFU counts in device regions were substantially lower than in paired stool samples across both subjects, despite variation along the length of the device (**Fig. 5c**). Similarly, the fraction of live cells varied substantially across regions, indicating localized differences in microbial viability within individual devices (**Fig. 5f**). Principal Coordinates Analysis (PCoA) of unweighted UniFrac distances showed the expected separation of saliva and stool samples, with CapScan-derived samples occupying intermediate positions between these sample types (**Fig. 5e**). Sectioning additionally revealed clustering of device regions by subject, indicating that inter-individual variation remained detectable despite strong spatial gradients along the intestinal tract.

**Figure 5:**
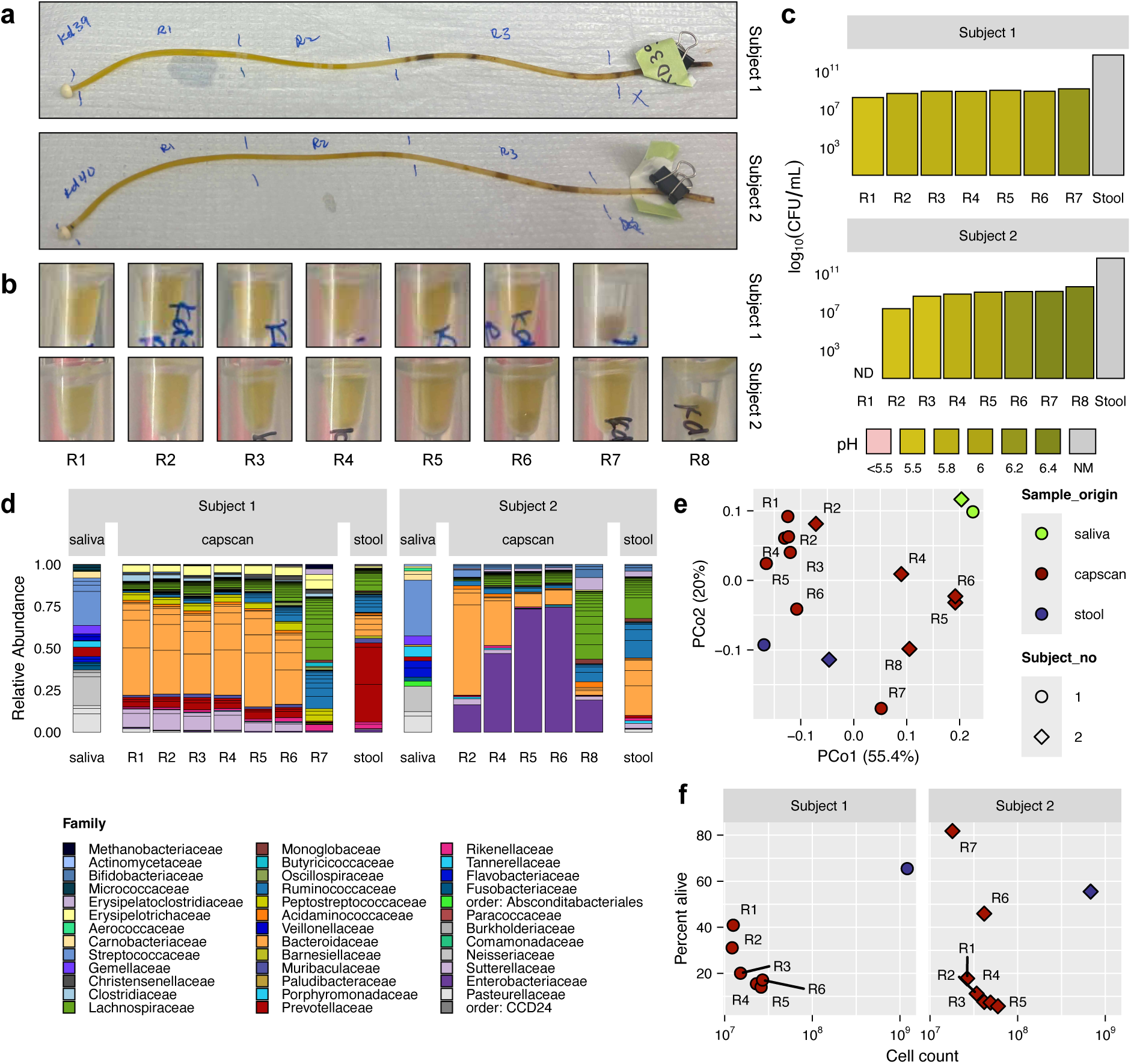
Spatial resolution along a CapScan Gen2 device. a) Photos of processed devices with annotations indicating how the device would normally be sectioned into three regions. Each device was instead sectioned into multiple sequential regions of ∼75 µL (3 drops) each and aliquoted for sequencing, CFU quantification, pH measurement, and microscopy-based total and live/dead cell counts. Shown are one device from each of two subjects. Paired saliva and stool samples were also collected (not shown). b) Photos of final collected sample aliquots. Note the darker color and lower sample volume of R7 for subject 1 and R8 for subject 2. c) CFU counts for each device region and paired stool sample for each subject. The CFU count for R1 from subject 2 was below the plating detection limit. Plating was performed on BHI–5% blood agar. Bars are colored by sample pH. pH measurements were obtained using pH indicator strips. When a pH range was assigned (e.g., pH∼5.8–6.0), the mean value is shown. d) Microbial composition of human saliva, SI, and stool samples. Stacked bars show ASVs with relative abundance >1%, colored by taxonomic family. Data are from 16S rRNA gene amplicon sequencing. Only samples with >400 reads are shown. e) Principal Coordinates Analysis based on weighted UniFrac distance shows clustering based on region as well as subject. f) Total cell counts and fraction of living cells obtained from single-cell epifluorescence imaging.

Although fine-scale gradients observed in several assays, the dominant spatial patterns were largely preserved when neighboring regions were consolidated, suggesting that three-region segmentation represents a practical balance between spatial resolution and sample volume for downstream analyses. Higher-resolution sectioning may nonetheless be advantageous when interrogating sharp metabolic transitions, localized host immune responses, or device-specific performance characteristics. Investigators should therefore align segmentation resolution with study objectives and available material.

## Methods

### Sample collection and processing

Human samples were collected under a protocol approved by the Stanford University Institutional Review Board (IRB #60005). Two subjects each ingested one CapScan Gen2 device approximately 3 h post-dinner and provided paired saliva samples at device ingestion and stool samples at device recovery. Within 1 h of device recovery, samples were transferred into an anaerobic chamber and processed as described in this Protocol. To increase spatial resolution, approximately 75 µL (∼3 drops) of CapScan Gen2 device contents were collected from sequential device regions for downstream analysis.

### Colony-forming unit (CFU) enumeration

For viable cell quantification, 1 µL of undiluted CapScan Gen2 sample or 1 µL of stool diluted 1:10,000 in 1X PBS was spread-plated onto BHI–5% blood agar. Plates were incubated anaerobically at 37 °C overnight before colony enumeration. CFU counts were calculated after accounting for dilution factors and plating volume.

### Cell quantification using microscopy

CapScan Gen2 samples were homogenized and imaged either undiluted or after a 1:100 dilution in 1X PBS, while stool samples were diluted 1:1,000. For each sample, 1 µL was spotted onto an agarose pad and imaged using phase-contrast and epifluorescence microscopy with a 100X oil immersion objective. Cells were stained with DAPI and propidium iodide (PI), or with the Live/Dead stain containing SYTO 9 and PI, to distinguish cells from food particles and debris. Dyes were added to the agarose pad before solidification. Images were acquired along a radial transect across each sample spot on the agarose pad to minimize spatial bias due to the coffee-ring effect. Cell density was quantified by automated segmentation using DeepCell^37^, a machine-learning-based image segmentation tool, and custom MATLAB scripts for post-processing. When live/dead staining was performed, cells were classified as live (SYTO 9 only), dead (PI only), or damaged (both SYTO 9 and PI). Percent viability was calculated as follows: % alive = [live / (live + dead + damaged)] × 100. This approach provided reproducible estimates of total cell density and viability under standardized acquisition parameters.

### Metagenomics and DNA concentration spike-in

Genomic DNA was extracted from samples using the DNeasy UltraClean Microbial Kit (Qiagen, cat. #10196-4) according to the manufacturer’s protocol. DNA concentrations were quantified using a PicoGreen dsDNA quantitation kit (ThermoFisher) and normalized prior to library preparation.

To estimate absolute microbial abundance, ZymoBIOMICS Microbial Community DNA Standard (Zymo Research) was spiked into normalized DNA samples following the manufacturer’s protocol. Shotgun metagenomic libraries were prepared using the Nextera XT kit (Illumina, cat. #FC-131-1096), pooled at equal volumes, and purified twice using AMPure XP beads. Libraries were sequenced on an Illumina NovaSeq platform with paired-end 150-bp reads. Illumina adapter sequences were removed using Skewer v. 0.2.2^38^. Absolute microbial abundance estimates were calculated based on spike-in DNA recovery and the known input concentrations of the ZymoBIOMICS standard.

### 16S rRNA gene sequencing and analysis

Genomic DNA was extracted from saliva, CapScan Gen2 device intestinal contents, and stool samples using the DNeasy UltraClean Microbial Kit (Qiagen, cat. #10196-4) according to the manufacturer’s protocol.

Libraries were prepared using a two-step dual-indexed barcoding strategy (CUPID-seq) with AccuStart II PCR SuperMix (Quantabio, cat. #95137-100). In the first PCR (27 amplification cycles), the V4 region of the 16S rRNA gene was amplified using Earth Microbiome Project primers 515F and 806R, modified with phase offsets to enhance base diversity. The second PCR (10 amplificatiion cycles) appended Illumina sequencing adapters and sample-specific barcodes. PCR products (5 µL per sample) were directly pooled, then purified and concentrated using the NucleoSpin Gel and PCR Clean-up Mini Kit (Macherey-Nagel, Fisher cat. #740609). The final library was sequenced on an Illumina NextSeq 2000 using P2 reagents with paired-end 150-bp reads.

Data were demultiplexed using UMI-tools v. 1.1.1^39^, and primer and adapter sequences were trimmed using cutadapt v. 1.18^40^. Quality filtering, trimming, denoising, and ASV inference were performed using DADA2^41^ as previously described^42^. Taxonomic assignment of ASVs was conducted using the SILVA reference database (nr99, version 138.2)^43^ with the assignTaxonomy() and addSpecies() functions in DADA2^41^.

## Data availability

All sequencing data have been deposited in the NCBI Sequencing Read Archive (SRA) under Project PRJNA1465842.

## Code availability

All code required for reproducing the analyses and figures are available at https://github.com/fubeverly/gen2-protocol.

## Author contributions statements

B.F., L.B.D., J.S., S.M., B.K., P.O., S.P.S., T.F., M.R., V.E.K., J.A.G., and S.E. carried out the experimental work and wrote the protocol. B.F. and S.E. wrote the original manuscript. S.S., D.A.R., J.A.G., D.S., S.E., and K.C.H. supervised the work. All authors contributed to the review and editing of this manuscript and approved the final manuscript.

## Acknowledgments

This work was supported by the National Science Foundation Postdoctoral Research Fellowship in Biology grant 2508176 (to B.F.) and National Institutes of Health grants T32 AI007328 (to B.F.) and DP1 DK147449 (to K.C.H.). K.C.H. is a Chan Zuckerberg Biohub Investigator. A subset of the figure icons were created with BioRender.

$Supplementary Tables

---

**Table S1:**
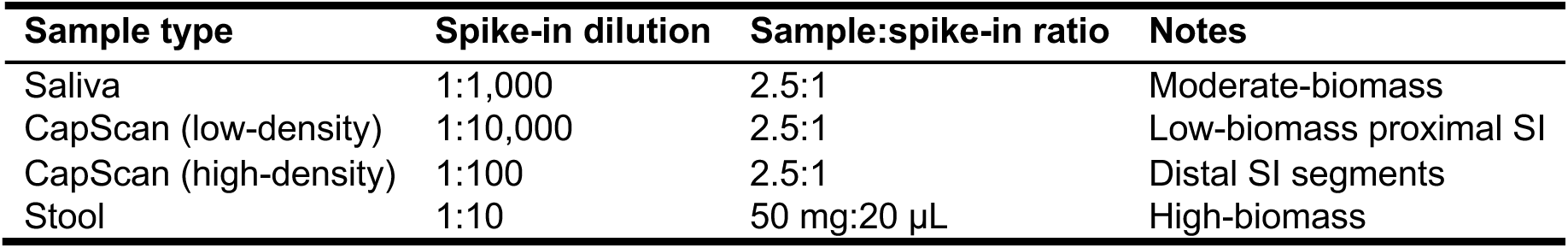
Recommended spike-in dilutions for absolute microbial quantification across sample types. Spike-in dilution refers to dilution of ZymoBIOMICS™ Spike-in Control I (High Microbial Load) in molecular-grade water prior to addition to sample. Sample:spike-in ratio indicates mixing ratio before DNA extraction. Dilutions should be empirically optimized based on expected biomass.

**Table S2:**
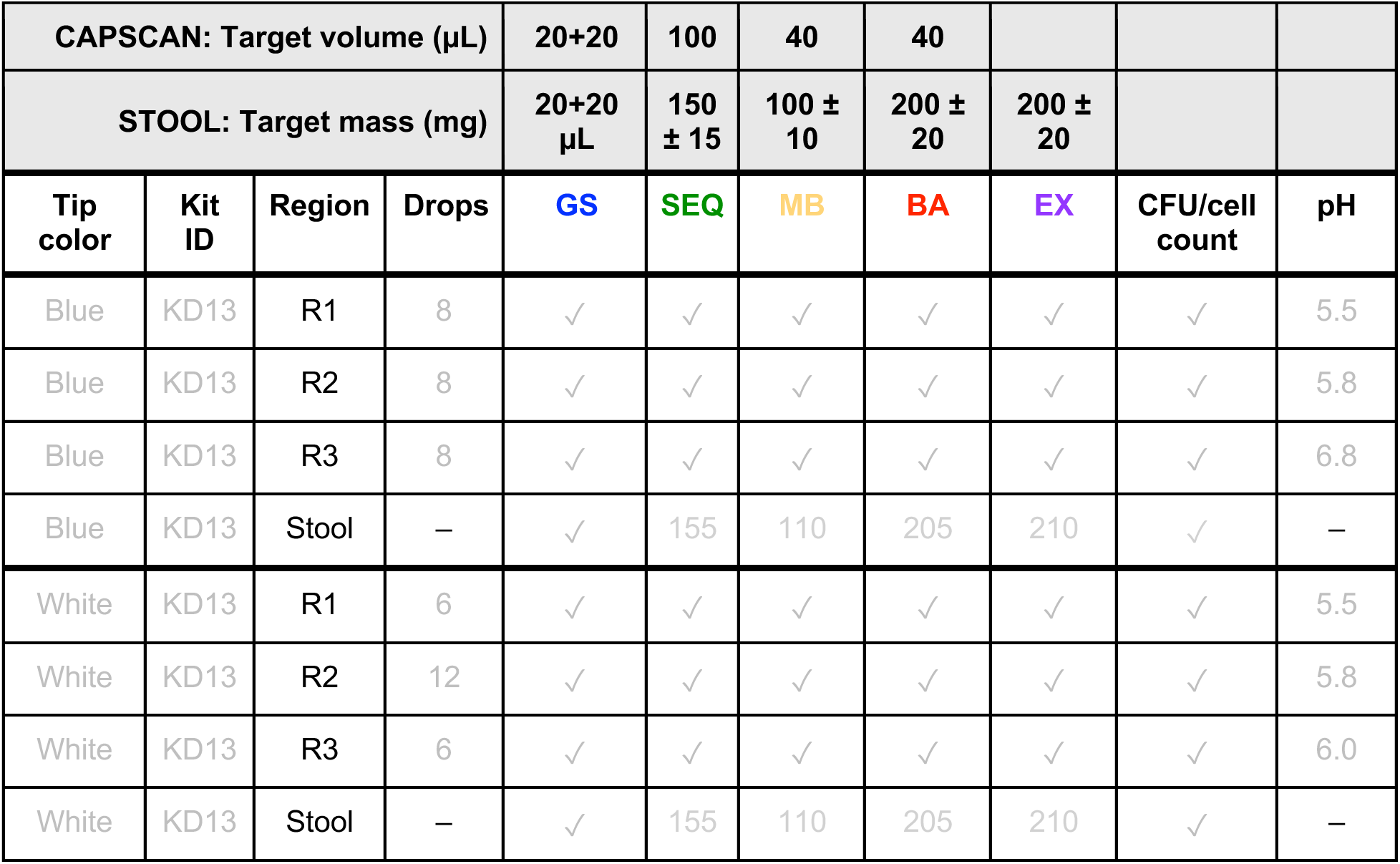
Example sample collection metadata record form.

| CAPSCAN: Target volume (µL) |  |  |  | 20+20 | 100 | 40 | 40 |  |  |  |
| --- | --- | --- | --- | --- | --- | --- | --- | --- | --- | --- |
| STOOL: Target mass (mg) |  |  |  | 20+20<br>µL | 150<br>± 15 | 100 ±<br>10 | 200 ±<br>20 | 200 ±<br>20 |  |  |
| Tip<br>color | Kit<br>ID | Region | Drops | GS | SEQ | MB | BA | EX | CFU/cell<br>count | pH |
| Blue | KD13 | R1 | 8 | ✓ | ✓ | ✓ | ✓ | ✓ | ✓ | 5.5 |
| Blue | KD13 | R2 | 8 | ✓ | ✓ | ✓ | ✓ | ✓ | ✓ | 5.8 |
| Blue | KD13 | R3 | 8 | ✓ | ✓ | ✓ | ✓ | ✓ | ✓ | 6.8 |
| Blue | KD13 | Stool | – | ✓ | 155 | 110 | 205 | 210 | ✓ | – |
| White | KD13 | R1 | 6 | ✓ | ✓ | ✓ | ✓ | ✓ | ✓ | 5.5 |
| White | KD13 | R2 | 12 | ✓ | ✓ | ✓ | ✓ | ✓ | ✓ | 5.8 |
| White | KD13 | R3 | 6 | ✓ | ✓ | ✓ | ✓ | ✓ | ✓ | 6.0 |
| White | KD13 | Stool | – | ✓ | 155 | 110 | 205 | 210 | ✓ | – |

